# Simple Accurate Regression-Based Forecasting of Intensive Care Unit Admissions due to COVID-19 in Ontario, Canada

**DOI:** 10.1101/2020.11.16.20231399

**Authors:** David N. Fisman, Ashleigh R. Tuite

## Abstract

The pandemic caused by SARS-CoV-2 has proven challenging clinically, and at the population level, due to heterogeneity in both transmissibility and severity. Recent case incidence in Ontario, Canada (autumn 2020) has outstripped incidence in seen during the first (spring) pandemic wave; but has been associated with a lower incidence of intensive care unit (ICU) admissions and deaths. We hypothesized that differential ICU burden might be explained by increased testing volumes, as well as the shift in mean case age from older to younger. We constructed a negative binomial regression model using only three covariates, at a 2-week lag: log_10_(weekly cases); log_10_(weekly deaths); and mean weekly case age. This model reproduced observed ICU admission volumes, and demonstrated good preliminary predictive validity. Furthermore, when admissions were used in combination with ICU length of stay, our modeled estimates demonstrated excellent convergent validity with ICU occupancy data reported by the Canadian Institute for Health Information. Our approach needs external validation in other settings and at larger and smaller geographic scales, but appears to be a useful short-term forecasting tool for ICU resource demand; we also demonstrate that the virulence of SARS-CoV-2 infection has not meaningfully changed in Ontario between the first and second waves, but the demographics of those infected, and the fraction of cases identified, have.

## Background

The pandemic caused by SARS-CoV-2 has proven challenging clinically, and at the population level, due to heterogeneity in both transmissibility and severity. Critical care resources have been recognized from early in the pandemic as an important societal chokepoint, and saturation of critical care capacity can result in catastrophic care outcomes (1). Subsequent to the spring 2020 wave of COVID-19, the Canadian province of Ontario saw a decline in daily cases and deaths, and reduced ICU occupancy. Since September 2020, exponential growth in cases has occurred again, with daily case counts exceeding those seen in spring, with (to date) fewer fatalities, and fewer individuals requiring intensive care (2). We noted that the age distribution of COVID-19 cases in Ontario had changed markedly since the spring wave, and also that the province was performing far more testing, suggesting that the apparent decrease in virulence might simply reflect younger age distribution of more recent cases, combined with inflated case counts due to increased case identification.

## Objectives

We sought to determine whether average age of cases, in combination with case counts and test volumes, can explain the apparent decline in COVID-19 virulence in Ontario, Canada; and whether simple regression models based on these covariates might provide a parsimonious and accurate means of near-term forecasting of ICU demand in the province.

## Methods and Findings

This study was approved by the Research Ethics Board of the University of Toronto. We obtained COVID-related ICU admissions, newly reported case counts, average weekly case age, and testing volumes for the province of Ontario to October 30, 2020, using the Integrated Public Health Information System (iPHIS), and Ontario Laboratory Information System (OLIS), as described elsewhere (3). Lags in case and ICU admission reporting were corrected by dividing the counts in the last 14 days of the time series by fractional reporting, derived using Kaplan-Meier methods. We constructed negative binomial regression models that predicted weekly ICU admissions based on average case age, log_10_(weekly test volumes) and log_10_(weekly case counts) at 2-week lags based on an average lag between symptom onset and ICU admission of 9.5 days (IQR 5 to 11 days). Resultant model fit was excellent (**Figure 1a, Table 1**).

**Table 1.**
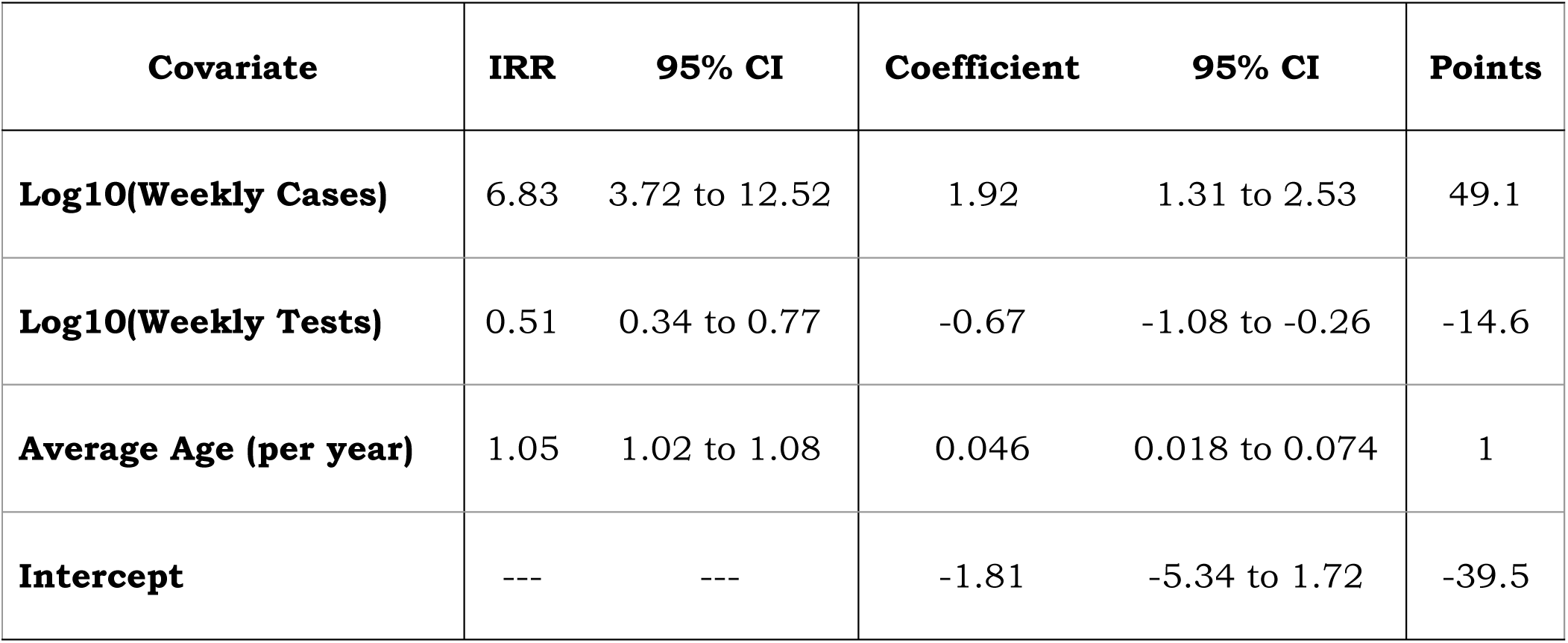
Regression Model Predicting Weekly ICU Admissions in Ontario, Canada at a 2-Week Lag.

| **Covariate** | **IRR** | **95% CI** | **Coefficient** | **95% CI** | **Points** |
| --- | --- | --- | --- | --- | --- |
| **Log10(Weekly Cases)** | 6.83 | 3.72 to 12.52 | 1.92 | 1.31 to 2.53 | 49.1 |
| **Log10(Weekly Tests)** | 0.51 | 0.34 to 0.77 | -0.67 | -1.08 to -0.26 | -14.6 |
| **Average Age (per year)** | 1.05 | 1.02 to 1.08 | 0.046 | 0.018 to 0.074 | 1 |
| **Intercept** | --- | --- | -1.81 | -5.34 to 1.72 | -39.5 |

**Figure 1.**
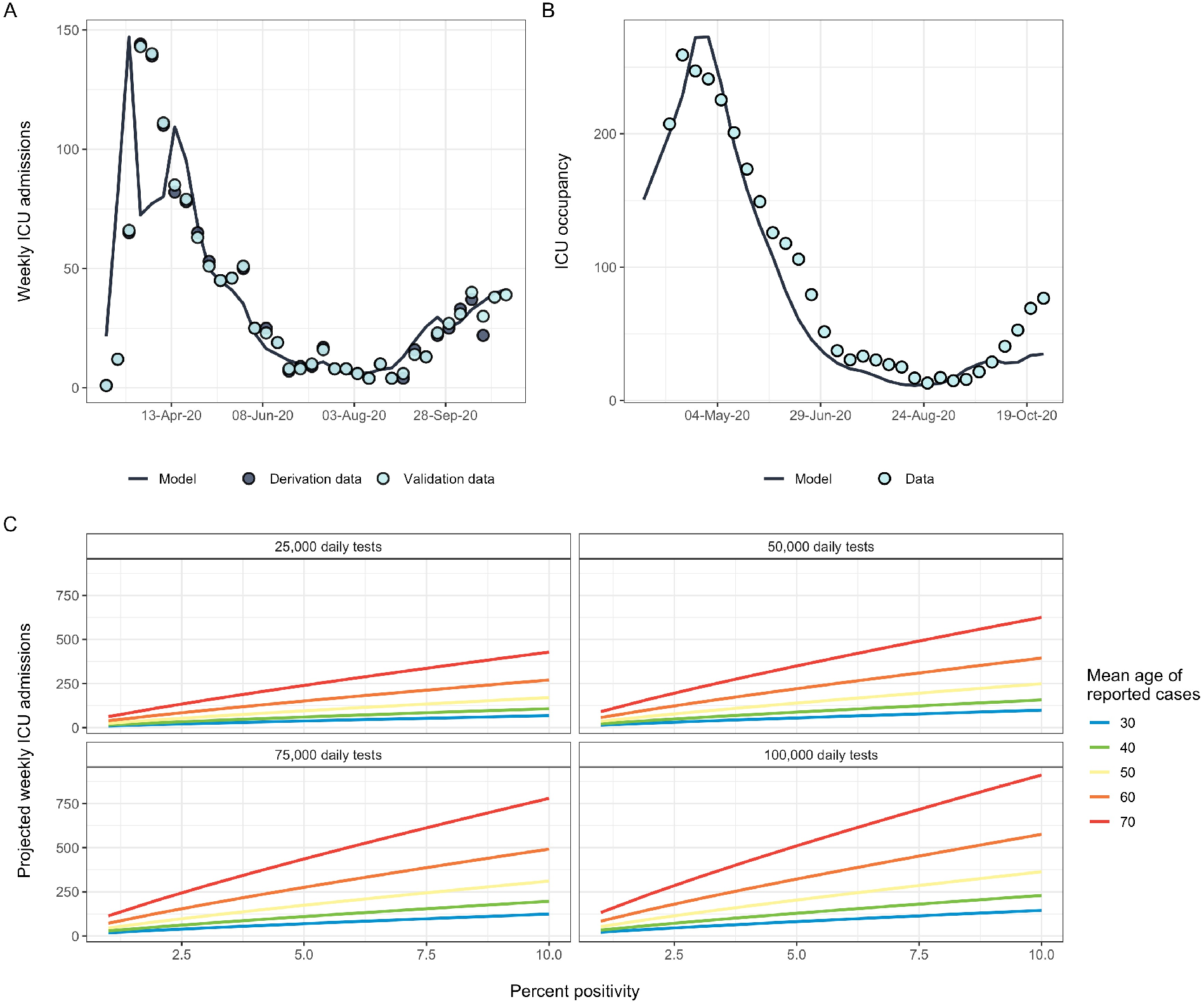
Regression Model Predicting Weekly ICU Admissions in Ontario, Canada at a 2-Week Lag. Panel (A) shows predicted weekly ICU admissions from a regression model (line) fit to data from an October 30, 2020 dataset (dark circles), with a later dataset (November 11, 2020) overlaid. Panel (B) shows excellent agreement beween model-based prediction of ICU occupancy and reported average ICU occupancy by week. Panel (C) demonstrates model-based projections of weekly ICU admissions (Y-axes) across a range of mean case ages, testing levels, and percent test positivity (X-axes).

We performed a preliminary evaluation of predictive validity using iPHIS and OLIS datasets from November 11, 2020. Running the model forward we saw good agreement between observations and expectations of ICU admissions. We assessed convergent validity of validated our ICU admission estimates by transforming them to ICU occupancy estimates by adding predicted admissions each week and removing current ICU occupants at a rate of 1/L, where L is expected average mean length of stay for individuals admitted to ICU in a given week. We compared these estimates to independent reports of COVID-19-related ICU occupancy in Ontario from the Canadian Institute for Health Information (CIHI); model-based estimates and CIHI reports for each week were well-correlated (intraclass correlation coefficient 0.98, 95% CI 0.97 to 0.99). (**Figure 1b**).

Our model covariates can be converted to a simple score: S = 49.1(log_10_(weekly cases)) + (mean age) – 14.6(log_10_(weekly tests)) – 39.5. Weekly predicted ICU admissions are then calculated e^(0.046S)^, where 0.046 is the smallest covariate from the negative binomial model. Predictions generated using such an approach, which can be used for planning purposes are presented in **Figure 1c**.

## Discussion

The marked variability in severity of SARS-CoV-2 infection by age makes surveillance of the pandemic using only case counts problematic. We show here that future ICU burden can be accurately forecast if case counts are used in conjunction with estimates of test frequency and mean case age. Others have previously noted that correction for test frequency is necessary for accurate forecasting and estimation of infection fatality (4, 5), and we have also noted previously that variability in population age structure is a key determinant of SARS-CoV-2 epidemic severity (6). Here we show that these elements considered in conjunction can provide a simple, apparently valid approach to near term forecasting of ICU utilization. A similar approach might be applied to mortality forecasting or hospitalization.

Limitations of this work include lack of prospective assessment of predictive validity, which is now ongoing, and also uncertainty around generalizability of this approach to other jurisdictions, and to larger and smaller geographic scales. Nonetheless, the apparent ability to accurately forecast ICU demand on a two-week time horizon may prove particularly useful to those charged with management of healthcare resource during the pandemic. Conceptually, our finding that case counts need to be presented in context of who (demographically) is infected and how many individuals were tested to arrive at a given case count, allows for more nuanced and meaningful risk communication during the current pandemic.

## Data Availability

Data available through Dr. Fisman.

## References

1. Lehmann E. Lessons from Northern Italy: Why even great Health Systems collapse under COVID-19 case load. Available via the Internet at https://globalhealth.harvard.edu/lessons-from-northern-italy-why-even-great-health-systems-collapse-under-covid-19-case-load/. Last accessed November 10, 2020. Harvard Global Health Institute; 2020.

2. Goodfield K. Ontario breaks another COVID-19 record with more than 1,300 new cases. Available via the Internet at https://toronto.ctvnews.ca/ontario-breaks-another-covid-19-record-with-more-than-1-300-new-cases-1.5179898. Last accessed November 9, 2020. CTV News; 2020.

3. Fisman DN, Greer AL, Hillmer M, O’’Brien S, Drews SJ, Tuite AR. COVID-19 Case-Age Distribution: Correction for Differential Testing by Age. medRxiv 2020.09.15.20193862; doi: https://doi.org/10.1101/2020.09.15.20193862. Available via the Internet at https://www.medrxiv.org/node/97162.external-links.html. Last accessed October 27, 2020.; 2020

4. Favero N. Adjusting confirmed COVID-19 case counts for testing volume. medRxiv 2020.06.26.20141135; doi: https://doi.org/10.1101/2020.06.26.20141135. Available via the Internet at https://www.medrxiv.org/content/10.1101/2020.06.26.20141135v1.full.pdf. Last accessed November 9, 2020. 2020.

5. Grewelle R, De Leo G. Estimating the Global Infection Fatality Rate of COVID-19. medRxiv 2020.05.11.20098780; doi: https://doi.org/10.1101/2020.05.11.20098780. 2020.

6. Fisman DN, Greer AL, Tuite AR. Age Is Just a Number: A Critically Important Number for COVID-19 Case Fatality. Ann Intern Med. 2020.

